# Impact of Clinical and Genomic Factors on SARS-CoV2 Disease Severity

**DOI:** 10.1101/2021.03.15.21253549

**Authors:** Sanjoy Dey, Aritra Bose, Prithwish Chakraborty, Mohamed Ghalwash, Aldo Guzman Saenz, Filippo Utro, Kenney Ng, Jianying Hu, Laxmi Parida, Daby Sow

**Affiliations:** Center for Computational Health, IBM Research, Yorktown Heights, NY, USA; Computational Genomics, IBM Research, Yorktown Heights, NY, USA

## Abstract

The SARS-CoV2 virus behind the COVID-19 pandemic is manifesting itself in different ways among infected people. While many are experiencing mild flue-like symptoms or are even remaining asymptomatic after infection, the virus has also led to serious complications, overloading ICUs while claiming more than 2.6 million lives world-wide. In this work, we apply AI methods to better understand factors that drive the severity of the disease. From the UK BioBank dataset we analyzed both clinical and genomic data of patients infected by this virus. Leveraging positive-unlabeled machine learning algorithms coupled with RubricOE, a state-of-the-art genomic analysis framework for genomic feature extraction, we propose severity prediction algorithms with high F_1_ score. Furthermore, we extracted insights on clinical and genomic factors driving the severity prediction. We also report on how these factors have evolved during the pandemic w.r.t. significant events such as the emergence of the B.1.1.7 SARS-CoV2 virus strain.

## Introduction

COVID-19, caused by the deadly “ severe acute respiratory syndrome coronavirus 2” (SARS-CoV2) virus, is one of the worst pandemic in human history inflicting severe casualties across the globe. So far, it has caused 117,332,262 confirmed cases while claiming 2,605,356 deaths worldwide^1^. This virus has stressed many health systems beyond their limits, screaming for an urgent reaction from research to help mitigate this unprecedented societal threat.

The outbreak of the virus first occurred in Wuhan province of China and then rapidly spread world-wide using human-to-human transmission^2^. One of the primary reason for the rapid spread is due to the wide spectrum of symptoms inhibited by the virus ranging from asymptomatic to mild to rapid progression to critical stage with pneumonia and likely death of respiratory failure^3^. In general, patients with mild symptoms cure easily whereas severe COVID-19 patients require further treatments such as ventilation in ICU. Such heterogeneity in terms of COVID-19 disease pose great challenges for designing treatment protocols. Typically, severe effects of infectious diseases like COVID-19 are hypothesized to be associated with the variations of host genome^4^. Several studies^5^ were conducted recently to find the biological mechanisms of the severity of COVID-19 diseases such as genome wide association studies (GWAS) of severe COVID-19^6^. In addition, clinical factors such as demographic, socio-economic factors, lifestyles, prior conditions may also have impact on the spread, exposure, and severity. So does the viral load of the SARS-CoV2 virus. The availability of rich electronic health records (EHR) provides an opportunity for finding such important clinical factors associated with COVID-19 along with important clinical factors observed in EHRs^3,7,8^.

Finding the bio-markers for severe COVID-19 patients requires rigorous study on clinical and genomic datasets. Most of the existing studies that aimed at finding the common risk factors for severe COVID-19 patients focused on either the clinical or the genomics factors, but not both. In this study, we aim to find the common risk factors associated COVID-19 severity using diverse factors from both clinical and genomics data available from a large-scale EHR dataset. Such kind of combined clinico-genomic factors can provide a holistic view of COVID-19 disease mechanisms. Specifically, it can provide the additional effects of genomic factors on the host genome as they relate to common clinical factors.

However, there are few challenges in finding the clinical and genomic factors associated with COVID-19 that can be used as bio-markers. *First*, defining the COVID-19 severity from EHR datasets is often challenging, since it is not collected directly in these datasets. Hence, the severity has to be defined using some clinical knowledge as a surrogate phenotype which in turn may introduce label noise and data collection bias. *Second*, assessing the impact of genomic factors on COVID-19 severity may be confounded by several other clinical factors such as the prior comorbidities of patients and treatment protocols. The effect of such confounding factors has to be addressed carefully when conducting the combined clinico-genomic studies. *Third*, the impact of clinico-genomic factors may also vary depending on the variations of diverse COVID-19 strains that have been observed due to genetic mutations.

In this study, we propose a combined framework for finding the clinical and genomic factors associated with COVID-19 severity using a large COVID-19 dataset from the UK Biobank (UKBB). To address the above mentioned challenges, we first use the COVID-19 related hospitalization as a surrogate outcome for defining severe COVID-19 cases. Second, we use a machine learning technique called positive-unlabeled (PU) learning to address the noise and reporting bias present in COVID-19 severity labels. Moreover, we use a recently proposed genomic analysis framework entitled RubricOE^9^ to select the set of genomic factors after adjusting for the common prior comorbid conditions which may act as potential confounders. Finally, we aim to assess how the importance of the extracted bio-markers evolve over the pandemic marked by event such as the emergence of the reportedly more contagious B.1.1.7 COVID-19 strain.

## Method

### Dataset

We analyzed the large prospective cohort of patients from the UK collected in the UKBB^10^ repository. The dataset contains diverse information including demographics, diagnosis, medications, lab tests, and genomic information of approximately half a million patients. In addition, the national SARS-CoV-2 laboratory test data were made available in UKBB through the Public Health England (PHE). This COVID-19 dataset contains a flag indicating *specimen origin* of COVID-19 tests: hospital inpatient origin vs other settings. It also notes the specimen collection date and the *specimen result* in addition to a few other information about how the specimen was collected. Data were available from March 16, 2020 and the repository has been updated once or twice every month. In our current study, we used data from March 16, 2020 to January 21, 2021.

### Preparing the outcome and feature sets

Although the COVID-19 dataset in UKBB is a prospective study, it did not collect the COVID-19 severity explicitly. The nature of this dataset is very diverse depending on national testing strategy which evolved over time as the pandemic progressed. For example, UK testing was initially restricted to those with symptoms in hospital and thus under this assumption, using the positive tests of these subjects in hospital can be a reasonable proxy for severity. However, when the testings were expanded covering even asymptomatic patients from diverse facilities, defining the severity required analyzing information regarding the specimen sources of test.

Among several different fields available in the COVID-19 table of our the UKBB, we settled on *specimen origin* and *specimen results* to track severity. We have used a combination of these flags for disease severity under the assumption that patients with a positive COVID-19 test obtained in hospital are likely to be severe cases. Table 1 contains the number of unique patients who had at least one positive test result and at least one ‘inpatient’ indicator retrieved from the origin field. We used inpatient samples COVID-19 patients (Origin=1 and result =1) as the severe COVID-19 patients while other COVID-19 patients (Origin=1 and result = 0) as the non-severe COVID-19 cases.

**Table 1:** The distribution of unique patients based on Origin and result as of Jan 21, 2021

|  | Result=0 | Result=1 | All |
| --- | --- | --- | --- |
| Origin=0 | 11,989 | 9,083 | 21,072 |
| Origin=1 | 31,800 | 4,345 | 36,145 |
| All | 43,789 | 13,428 | 57,217 |

While this approach estimates severity, it may be prone to a selection bias problem by defining all inpatient COVID-19 patients as severe cases. For instance, some patients may have been hospitalized due to other conditions or may have been experiencing other problems before they got the COVID-19 infection. To cope with aspects of this problem, a separate field covering hospitalization-acquired infections was collected in the database to filter out such patients from actual severe COVID-19 infection related hospitalization. However, this field was enabled in the dataset very recently and the protocol for marking that field is not yet uniformly applied to all samples.

Another potential issue with the dataset pertains to defining the non-severe COVID-19 patient cohort. While hospitalization data for acute COVID-19 patients were collected carefully, the non-severe patients are harder to define. For example, patients who were tested positively for COVID-19 in outpatients or non-urgent care service centers (Origin=0) may have experienced severe symptoms. The lone presence of the Origin flag set to 0 does not always mean that patients did not have severe reaction at a later stage - they may not have been traced prospectively in the dataset. Such tracing would most likely require conducting another test while being in an inpatient service. To cope with this issue, we used a special class of machine learning to handle this kind of noise associated in the outcome labels.

To build our severity prediction models, we used several clinical and genomic features that may have an impact on COVID-19 severity. For the clinical features, we manually curated some of the most relevant features which may have association with COVID-19 for further analysis. Our clinical features include demographics, lifestyles, comorbidities based on patient’s prior history, self-reports, and their chronic disease histories curated from prior inpatient records.

### Selecting Genomic Features

Starting with 12,965 overlapping samples with genomic information in the UKBB COVID-19 dataset, we only retained high quality imputed genomic markers in the form of Single Nucleotide Polymorphism (SNPs) with imputation quality INFO score *>* 0.3, resulting in approximately 4.4 million SNPs. We further did a standard quality control (QC) for filtering informative variants, removing SNPs as well as individuals with more than 2% of missing data. We further filtered for Minor Allele Count (MAC) *>* 2, Minor Allele Frequency (MAF) *>* 0.05, sex discrepancy in individuals, variants deviating by more than *p <* 1*e —* 6 in the Hardy-Weinberg equilibrium (HWE) for cases and *p <* 1*e −*10 in controls. We also filtered for individuals with *±*3 standard deviation in heterozygosity rates and very closely related individuals (second degree). All of the QC analysis was done using PLINK v1.9^11^. We ended up with 12,389 individuals and 4,539,795 SNPs which passed all the QC thresholds. Of these 12,389 individuals, 4,000 were cases with severe COVID-19 infection and 8,389 were controls or non-severe individuals who tested positive. We further pruned for linkage disequilibrium (LD), removing correlated SNPs with *r*^2^ *>* 0.2 with a variance inflation factor of 10 on a window size of 50 kb. We used this pruned dataset for Principal Component Analysis (PCA) and applying downstream algorithms to select stable features.

### Genome-wide association study

We performed a GWAS on the filtered dataset to perform a sanity check for the QC analysis as well as observe significant loci associated with the severity of COVID-19 infection. GWAS was performed by the package SAIGE^12^, which accounts for case-control imbalance in the data. We performed genome-wide association tests while correcting for known confounders in genomics such as population structure, sex and age. We computed the top 20 principal components (PCs) with TeraPCA^13^ as a proxy for population structure and used them as covariates along with the biomarkers^14^.

### Stable feature selection

We applied RubricOE (learning rubric for multi-omics and genetic epidemiology), a framework for learning stable features discriminating cases from controls to understand significant SNPs associate with COVID-19 severity. The rubric employs a nested test-training set configuration on the genomic data after QC and pruning for LD. The outer test-training set split reserves the test set (“ validation”) for final SNP evaluation and denotes the training set as “ working” data. Within the latter, further train-test splits are performed to rank the features by their Youden index^15^ (also known as J statistic). The pipeline of RubricOE is outlined in Figure 1. RubricOE is applied on the genomic data, accounting for top 20 PCs, sex, year of birth and Diabetes Mellitus (DM) as covariates. It iteratively finds a “ stable” set of SNPs which persistently remains highly ranked with their respective Youden index.

**Figure 1:**
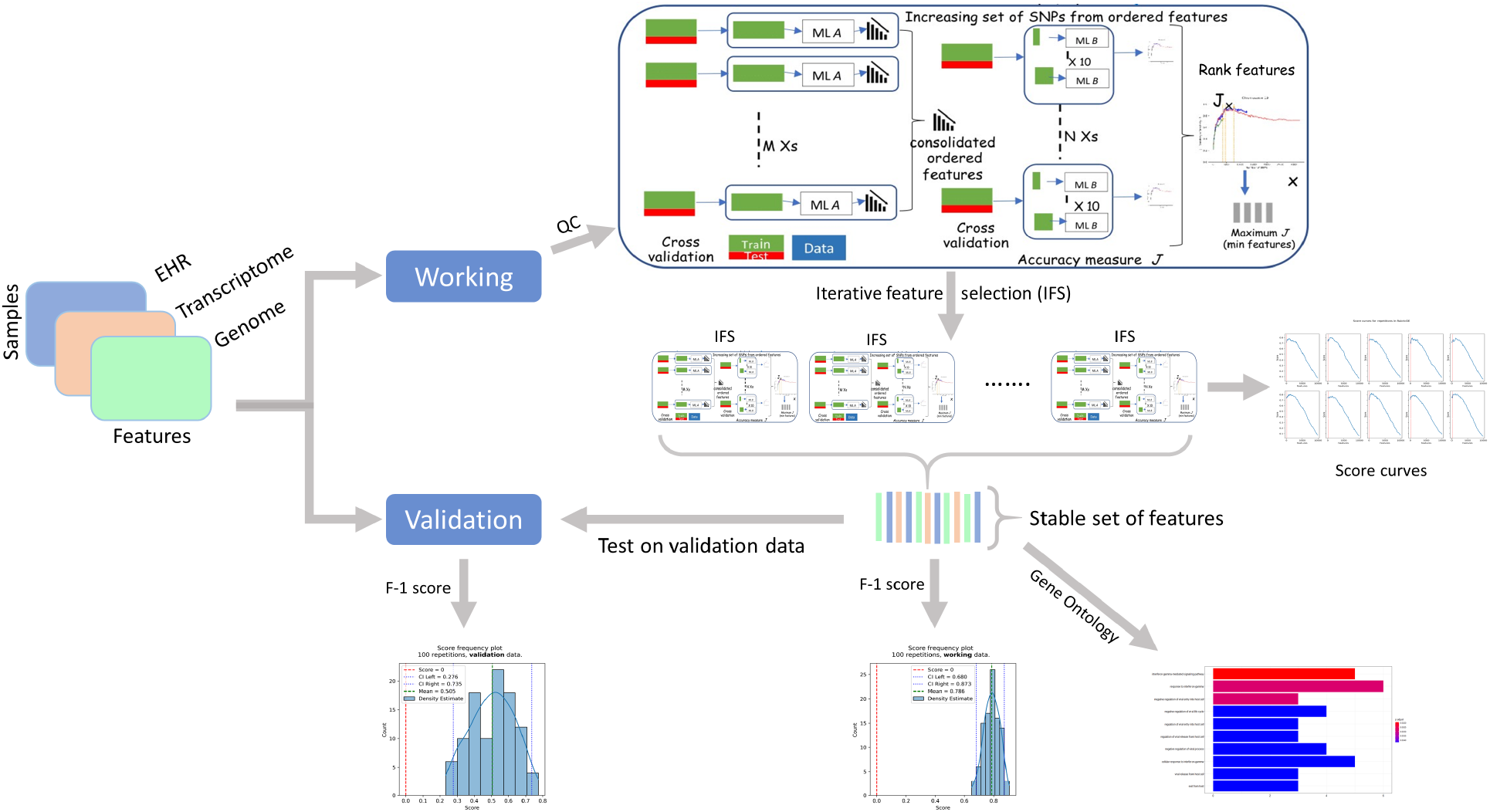
Components of RubricOE, as ensemble ML pipeline extracting stable features from multi-omics data.

### Functional annotation

Annotation, Gene Ontology (GO) and enriched pathway analysis of the “ stable” set of SNPs produced by RubricOE has been evaluated using the R package clusterProfiler^16^ v3.12.

### Framework

We used a predictive learning framework called Positive-unlabeled learning (PU learning) for extracting the most significant clinical and genomic features associated with COVID-19 outcome. Note that our dataset is more robust in terms of defining the severe COVID-19 patients than the non-severe cases. Hence, using a classical binary classification algorithm will not be appropriate and able to cope with such noisy class labels. PU learning algorithms have alternatively been reported to be especially suitable for learning from a noisy negative class by treating instances in this class as unlabeled while predicting the positive class (severe COVID-19 cases).

PU learning is a variant of the classical binary classification problem, where it is assumed that the unlabeled examples could belong to either positive or negative class. We refer to^17^ for a comprehensive review of such PU learning techniques. Among several state-of-the-art PU learning techniques, we use PU Adapter^18^, uPU^19^, and nnPU^20^ for learning the COVID-19 severity. All of these PU learning techniques have the basic assumption that the observed samples are drawn uniformly from the positive distribution, which enables PU learning techniques to leverage standard binary classification methods with minor modification of data or algorithm.

The PU-Adapter^18^ technique pre-processes the available PU data so that any standard binary classification method can be used for learning on the PU dataset. In particular, this method reweighs the samples of PU data so that learning a traditional binary classifier on the weighted samples yield to similar target probability threshold on the PU dataset. Moreover, it has been shown that the classifier trained on positive and unlabeled examples predicts probabilities that differ by a constant factor from the true probabilities of positive class. Unbiased PU learning (uPU)^19^ uses two separate loss functions for the positive and unlabeled samples using convex optimization framework.

### Evaluation

We evaluate the effectiveness of PU-learning algorithms with a comparison with baseline traditional classification algorithm using logistic regression with a *L*_1_ loss. Computing traditional metrics such as precision, recall, *F*_1_ score, and accuracy is challenging in the positive unlabeled scenario since the only information available relates to the positive label while no sample from the negative class is clearly provided. One straightforward solution to this problem is to assume that the unlabeled data are negative. However, this is not fully accurate. A modified *F*_1_ score (we call it as 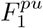 score) has been proposed to address this issue^21^. This score is computed as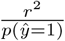, where *r* is the recall and *p*(ŷ = 1) is the prevalence of the predicted positive label. As shown in^21^, the 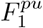 score can take values greater than one but shares the same property as *F*_1_ score; it is high when both precision and recall are high and small when either one is small. We also report metrics used to evaluate traditional classification methods for completeness although these are not recommended for our setting. They include accuracy, precision, recall, and *F*_1_ score.

We used a 5-fold cross-validation (CV) approach for evaluating the performances of the predictive models, where in each CV fold, the hyper-parameter for *L*_1_ loss was tuned internally using a grid-search over the range of [10^*−*3^, 10^*−*2^, 10^*−*1^, 1, 10]. Finally, we used a bootstrapping technique for estimating the confidence intervals of the scores using 100 bootstrapping runs for finding the importance of the clinico-genomic features.

## Results

The results of the baseline *L*_1_ regularized logistic regression are shown in Table 2. We report the metrics for three different models: clinical, genomic, and the combined clinico-genomic model. Overall, we can observe that clinical model has worse predictive power than the model built from genomic features obtained from GWA data. However, the combined model has significant improvement in terms of predictive power.

**Table 2:**
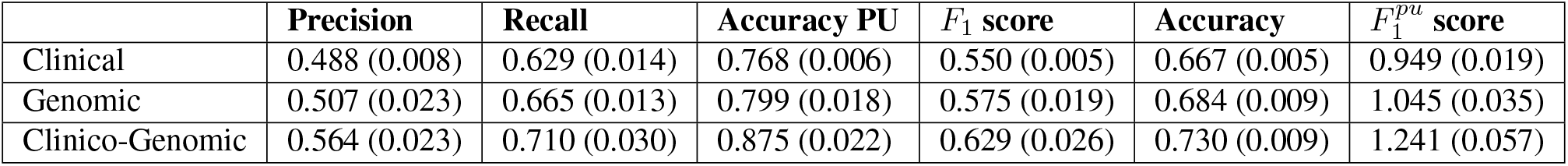
Average (standard deviation) 5-fold CV performances of three models based on Logistic Regression with *L*_1_ loss

Table 3 and Table 4 show the same metrics from the three different models using two different PU learning techniques: nnPU and PU-Adapted logistic regression, respectively. The nnPU model does not perform well for categorical genomic features, moreover it is harder to interpret such model. In contrast, the PU-adapted method uses logistic regression as the baseline model. Hence it is easier to interpret the resulting coefficients as log-odds ratios of clinico-genomic features. The combined clinico-genomic model of the PU adapted logistic regression model yields significant improvement from the models built on clinical and genomic model individually. Although the *F*_1_ score of the PU learning model is similar to the score of the baseline logistic regression model, the adjusted *F*_1_ score is higher for the PU model, which justifies the use of the PU model for our noisy class level.

**Table 3:** Average (standard deviation) 5-fold CV performances of three models based on nnPU Model

| | **Precision** | **Recall** | **Accuracy PU** | $F_1$ **score** | **Accuracy** | $F_1^{pu}$ **score** |
| --- | --- | --- | --- | --- | --- | --- |
| Clinical | 0.586 (0.017) | 0.416 (0.019) | 0.803 (0.003) | 0.486 (0.009) | 0.716 (0.006) | 0.754 (0.022) |
| Genomic | 0.4 (0.545) | 0 (0.000) | 0.677 (0) | 0.001 (0.001) | 0.677 (0.000) | 0.004 (0) |
| Clinico-Genomic | 0.830 (0.025) | 0.054 (0.010) | 0.694 (0.003) | 0.100 (0.018) | 0.691 (0.003) | 0.138 (0.027) |

**Table 4:** Average (standard deviation) 5-fold CV performances of three models based on PU Adapted Logistic regression
Learning

| | **Precision** | **Recall** | **Accuracy PU** | $F_1$ **score** | **Accuracy** | $F_1^{pu}$ **score** |
| --- | --- | --- | --- | --- | --- | --- |
| Clinical | 0.401 (0.008) | 0.800 (0.011) | 0.621 (0.022) | 0.534 (0.006) | 0.548 (0.015) | 0.991 (0.028) |
| Genomic | 0.417 (0.004) | 0.862 (0.006) | 0.660 (0.010) | 0.562 (0.005) | 0.567 (0.007) | 1.114 (0.018) |
| Clinico-Genomic | 0.485 (0.006) | 0.839 (0.021) | 0.794 (0.012) | 0.615 (0.009) | 0.660 (0.007) | 1.260 (0.040) |

We interpret the log-odds of the PU-Adapted logistic regression model to denote the importance of a particular feature toward COVID-19 severity. Figure 2 shows the feature importance of top 50 statistically significant features associated with COVID-19 severity computed by the final combined clinico-genomic model using the PU learning framework. The statistical significance of a feature is computed using bootstrapping with 100 runs and the confidence interval is represented as error bars in the figure which are beyond zero (the vertical line).

**Figure 2:**
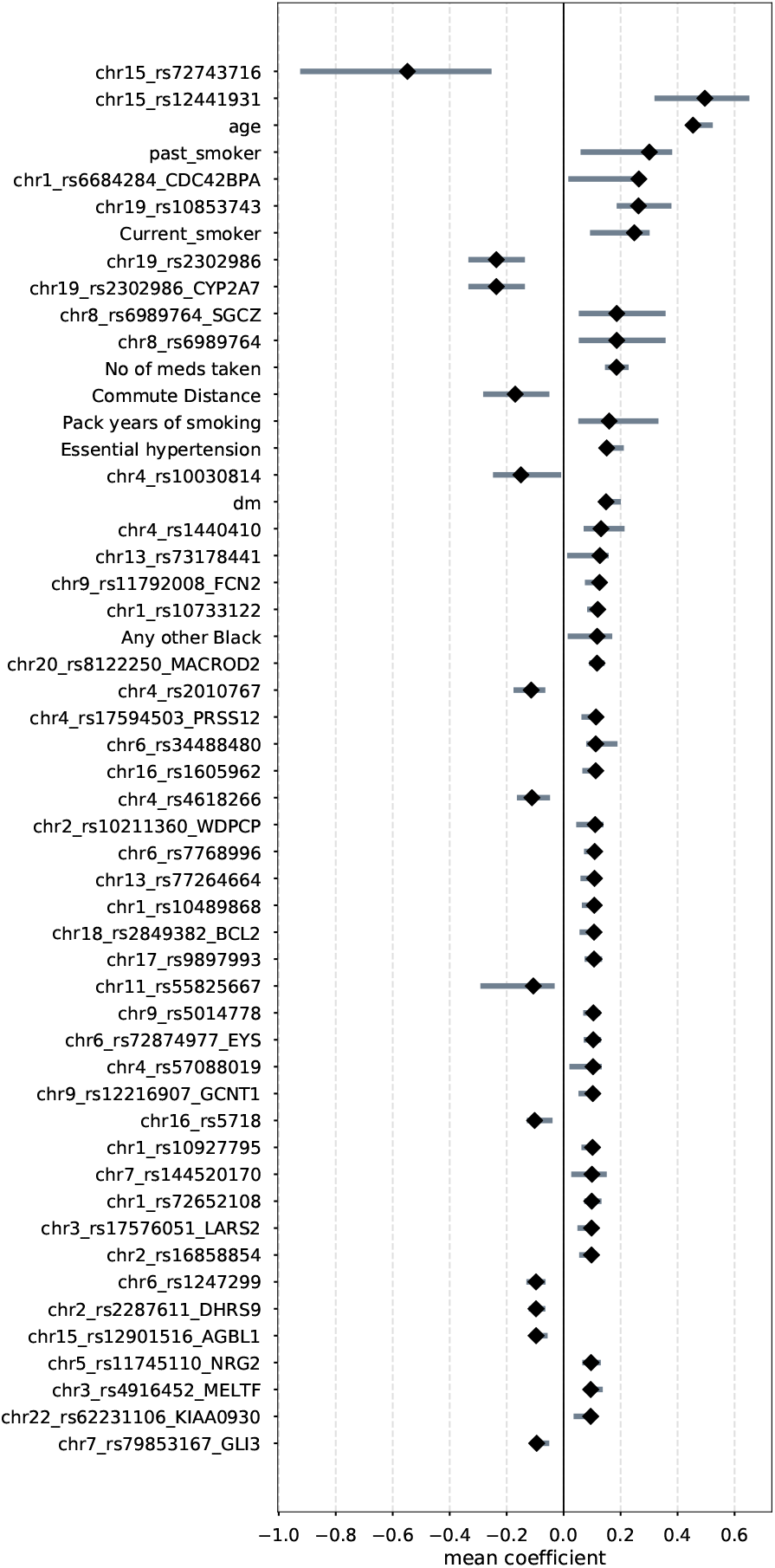
Top statistically significant features obtained from PUAdapted Logistic Regression. Plots show mean coefficients (log odds ratio) for regression along with 95% bootstrapped confidence interval.

## Discussion

Our final combined clinico-genomic model finds the significant features associated with COVID-19 severity (Figure 2), which may act as potential biomarkers. Some of the top significant clinical features have already been found in previous studies. For example, age and black ethnicity have been reported in^8^, and prior conditions like diabetes mellitus, hypertension have been reported in^7,22,23^. Note that our study finds smoking status (past smoker, current smoker, pack years of smoking) is related to severe COVID-19 patients, which has ambiguous evidence in literature. A few studies^8^ reported it with increased risk factors, while others^22^ did not find it to be a significant risk factors for COVID-19 severity.

In addition, our study found a few more features such as number of medication taken per day to be significant. Such features are related to the overall health status of the patient and correlate indirectly to high severity risk factors. Interestingly, distance from home to workplace has also been reported with lower risk of severity, which may be confounded by people commuting by car for longer distance instead of public transport or cycling for shorter commuting distance.

The genomic features obtained from RubricOE are enriched in a host of biological pathways related to transmembrane transporters and receptor activities (Figure 5(a)). The most significant is *transmembrane receptor protein tyrosine phosphatase* which on increasing activity might affect T cells and contribute to their depletion and immunoparalysis in severe COVID-19 patients^24^. Other important pathways such as *ion channel, metal ion* and *inorganic cation* transmembrane transporter activities reaffirm the notion that SARS-CoV-2 E protein is a potential ion channel^25^ like other coronaviridae^26^. Focusing on the significant biomarkers from the combined model, we observed two new GO enriched terms, *passive transmembrane transporter activity* and *PDZ domain binding* (Figure 5(b)). It has been shown that PDZ-containing proteins among binders of the SARS-CoV-2 proteins E, 3a or N affect viral replication under knock-down gene expression in infected cells, significantly^27^, particularly in the E protein^28^.

The impact of clinico-genomic features can be confounded by different treatment protocols. However, our UKBB dataset does not have direct information on COVID-19 drugs and detailed treatment protocols that a COVID-19 patient went through, so we tried to assess the impact of a COVID-19 drug indirectly using its date of approval. In particular, we assessed how the co-efficients of topmost features mentioned in Figure 2 change before and after the drug was approved for treatment. As a simple use-case, we report the changes of impact of the top 25 biomarkers (instead of all 50 due to space limitation) for the first approved UK drug called ‘Dexamethazone’ with its approval date as June 15, 2020 in Figure 3. We can observe that most of the 25 significant factors from the whole dataset still had similar impact after the drug being used, but only 5 features had significant impact before the drug was used.

**Figure 3:**
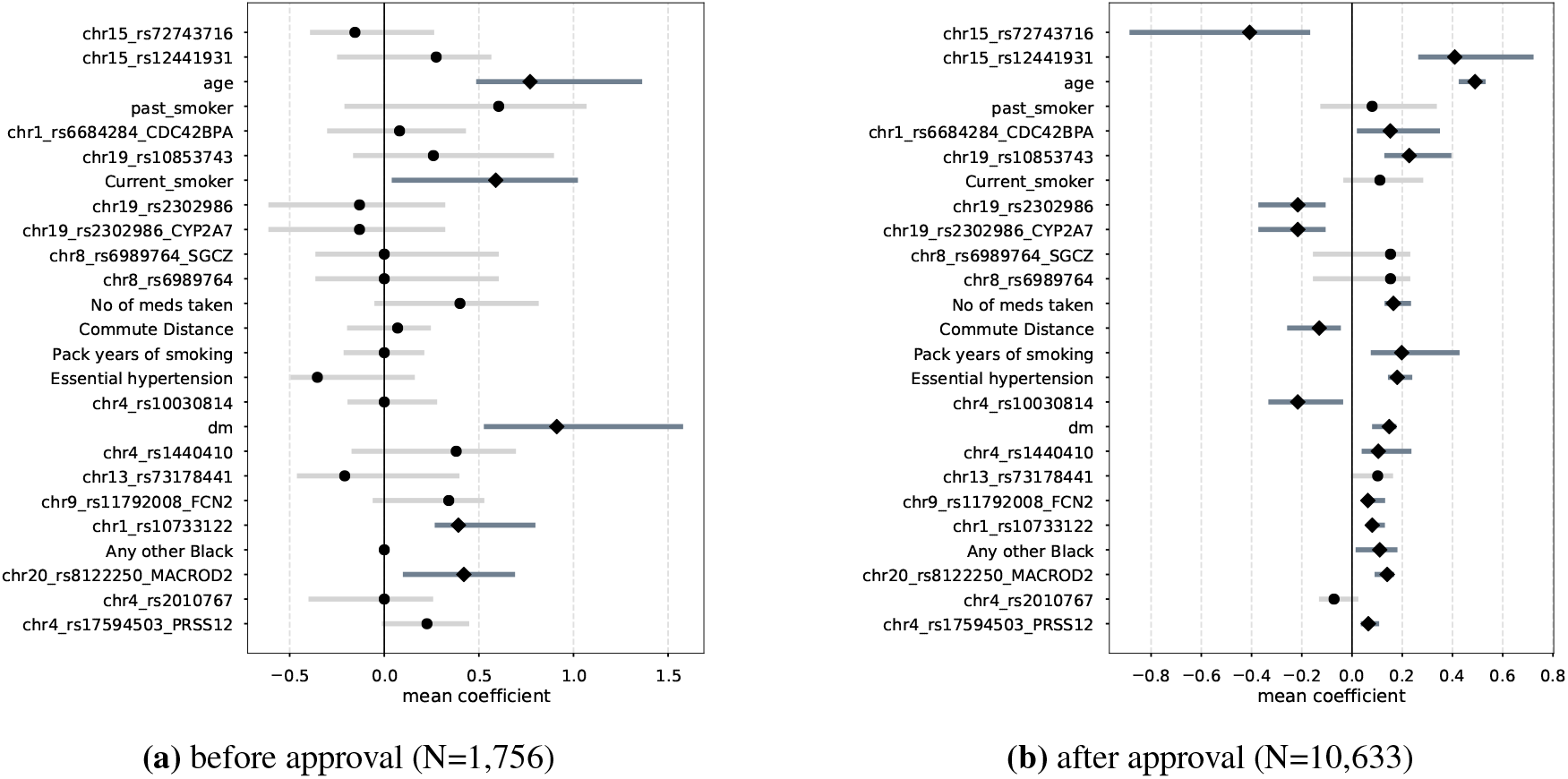
Mean coefficients with 95% CI of features from Fig 2 indicating severity of COVID-19 (a) before and (b) after Dexamethasone was approved for usage in UK. Features that are still significant (not significant) are shown via deep grey (light grey) bars with square (circular) markers.

Similarly, we also wanted to assess how the impacts of clinico-genomic features change over different strains of SARS-CoV-2. In the UK, a reportedly more contagious strain named B.1.1.7^29^ started surfacing from early September 2020 and it is estimated that by December 2020, over 60% of new COVID-19 patients had the newer strain. The UKBB also did not collect the virus genome prospectively. So the only way to assess the impact of clinico-genomic biomarkers is through the date when a patient was diagnosed with COVID-19. We divided the whole cohort into three parts: those diagnosed before Aug 31, 2020 (older strain), diagnosed between September 1st, 2020 and December 15, 2020 (both strains) and after December 15, 2020 (newer strain). Figure 4 shows how the effect of the top clinico-genomic features obtained by the global model change over these three cohorts. We can observe that some factors like age, prior DM condition and number of medication are consistent across both strains, while the smoking history (current and present) had larger impact on older strain than the newer strain. Such analysis of temporal trend of a factor’s impact on severity can help elucidate important clinical insights.

**Figure 4:**
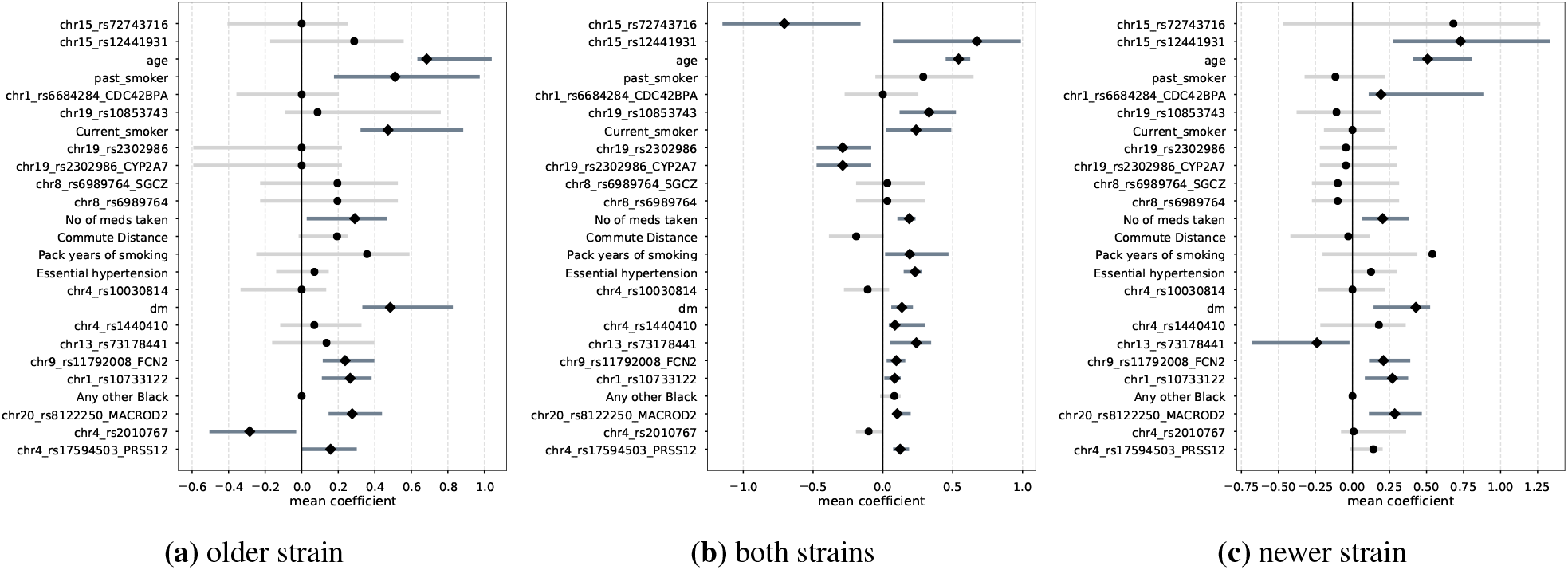
Mean coefficients with 95% CI of features from Fig 2 indicating severity of COVID-19 with dominance of (a) older strain, (b) both strains, and (c) newer strain (B.1.1.7) in UK. Features that are still significant (not significant) are shown via deep grey (light grey) bars with square (circular) markers.

**Figure 5:**
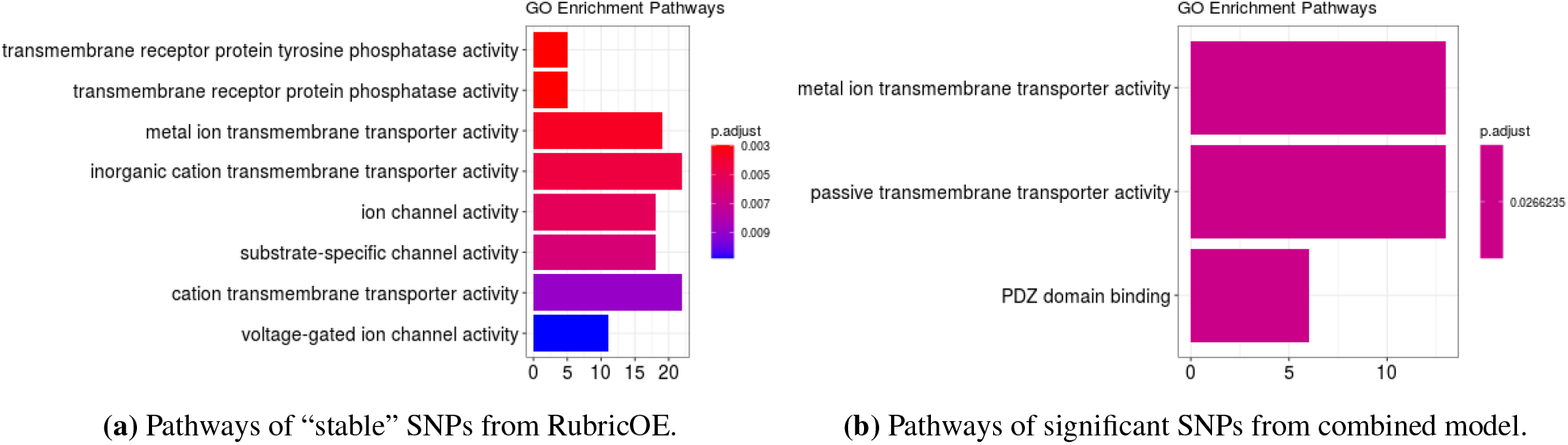
Boxplots of significant GO enriched pathways, colored by their corresponding corrected *p*-value and count of mapped genes in the x-axis.

## Related Works

To combat the global pandemic caused by COVID-19, scientists across different disciplines have been studying the disease to understand various facets such as its spread, epidemiological and patho-physiological characteristics, and societal impact. In this section we survey some of the most relevant literature.

Some of the more studied aspects of understanding the spread and clinical manifestations of the disease has been focused on the viral and host genomic factors^30^. To help consolidate and streamline the efforts around studying the host genetics for susceptibility the COVID-19 Host Initiative was launched in mid 2020^4^ with a primary focus on disease severity. As part of this initiative, Hou et al.^31^ found that ACE2 or TMPRSS2 DNA polymorphisms were likely associated with genetic susceptibility of COVID-19, while Li et al.^32^ found that genetic variants under a polygenic model shows promising improvements in prediction accuracies and argued for greater investigation into polygenic risk scores. Wang et al.^33^ presented further genetic insights into phenotypic difference among the COVID-19 patient groups, highlighting genes and variants of interest. More superficially, they conducted both single-variant and gene-based association with respect to five severity groups finding insights such as variants involved in IL-1 signaling pathways to be of interest. While studies identified a gene cluster on chromosome 3 as a risk locus for respiratory failure after infection with COVID-19, Zeberg et al.^34^ found that the risk is conferred by a genomic segment that is carried by around 50% of people in South Asia and 16% of people in Europe.

Simultaneously, there has been concerted efforts to characterize the clinical characteristics of COVID-19 including the patho-physiology and the risk factors for covid. Several large-scale meta-analysis studies were conducted either on previous studies (Fu et al.^3^) or by summarizing published articles (e.g., Li et al.^7^, Zheng et al.^8^) to identify common clinical symptoms and lab abnormalities among severe COVID-19 patients. Some common risk factors observed among critical/mortal COVID-19 patients are socio-demographic information such as male, older than 65, smoking, and prior history of hypertension, diabetes, cardiovascular disease, respiratory diseases, kidney diseases, obesity, immunosuppresion, and cancer. In addition to these prior conditions, depression as well as other cognitive and neurological disorders were also found as risk factors by a nationwide cohort study in Israel ^22^, although this study found that smoking and presence of respiratory diseases do not significantly increase the risk of complications. Williamson^23^ presented a risk analysis using a secure health analytics platform (OpenSAFELY) that covers 40% of all patients in England. They found risk factors such as male, age, socio-economic deprivation, diabetes, severe asthma, and various other medical conditions. They also found Black and South Asian people to be at higher risk. Research has also been conducted to characterize risk among sub-populations such as obesity, heart failure, and Parkinson’s Disease^35–37^ concurrently. Mathew et al.^38^ found three immunotypes revealing different patterns of lymphocyte responses among hospitalized COVID-19 patients.

Besides finding such clinico-genomic risk factors of severe COVID-19, several machine learning (ML) and artificial intelligence (AI) have also been applied successfully to solve a wide range of needs, ranging from fast ML based COVID-19 infection prediction ^39^ to routine blood-tests^40^ as fast alternatives to costly and time-consuming PCR tests^41^ to predicting disease state of individual cells given their transcriptomes^42^.

## Conclusion

In this study, we looked for significant clinical and genomic factors that are associated with severe reaction of SARS-CoV-2 virus infection from a large-scale EHR and GWA dataset available from the UK. In particular, we used a special class of machine learning called positive-unlabeled learning to address the challenge of noisy class label of COVID-19 severity outcome. Our holistic clinico-genomic modeling can potentially discover the effect of such factors in a robust manner, which can be used as potential biomarkers for better clinical decision making. In future, we plan to further investigate the effect of treatment protocols and vaccination on COVID-19 severity leveraging the inpatient data. Also, investigating risk factors associated with COVID-19 induced death will be an important future research.

## Data Availability

The data that support the findings of this study are available from the UK Biobank upon reasonable request. [1]
1. UK Biobank ACCESS PROCEDURES: Application and review procedures for access to the UK Biobank ResourcAccess procedures. Published 2011. https://www.ukbiobank.ac.uk/wp-content/uploads/2012/09/Access-Procedures-2011-1.pdf

https://www.ukbiobank.ac.uk

## Notes

### Competing Interest Statement

The authors have declared no competing interest.

### Funding Statement

Nothing to declare

### Author Declarations

Data analysis was performed under UK Biobank application 50658 using existing publicly available and deidentified data and was IRB exempt.

